# A preventive school-based paradigm using virtual reality technologies for improving emotional regulation, depressive and anxiety symptoms in children and adolescents (e-Emotio project): a randomized controlled pilot trial

**DOI:** 10.1101/2024.04.17.24305984

**Authors:** Anna Carballo-Marquez, Anna Garcia-Casanovas, Aikaterini Ampatzoglou, Juliana Rojas-Rincon, María Fernández-Capo, María Gámiz-Sanfeliu, Maite Garolera- Freixa, Bruno Porras-Garcia

## Abstract

**Introduction:** Preventive transdiagnostic interventions for depression and anxiety symptoms in children and adolescents are important in reducing the development of disorders later in life, and emotion regulation (ER) is one potentially relevant factor to consider. Impairments in executive functions play a critical role in emotion development and regulation in these ages. Immersive Virtual Reality (IVR) technology has exhibited potential to augment traditional cognitive training interventions. In this project, we propose to use Enhance IVR, a gamified cognitive training that can be tailored to specific ER strategies that individuals may face, such as difficulty with attention control, inhibition, or cognitive flexibility.

**Methods and analysis:** This is a longitudinal, parallel, single-blind, randomized, controlled pilot trial with an estimated sample of 160 participants. The first group (experimental group) will receive the gamified IVR program (Enhance IVR), while the second group (active control group) will receive a comparable IVR relaxation experience. Both Enhance IVR and the control IVR interventions will last five weeks, two times a week, 30 minutes (10 sessions per participant). Participants will undergo a baseline assessment that includes several measures related to mental health, ER, executive functioning, and cognitive performance, and another post intervention assessment with the same measures. Finally, variables related to system usability and cybersickness with the IVR tool will be evaluated for both groups.

**Ethics and dissemination:** This study was approved by the Ethical Committee of the International University of Catalonia (PSI-2023-04). The findings will be disseminated through peer-reviewed journals, reports, conferences, and other scientific events.

**Strengths and limitations of this study:**

- This is a controlled, randomized, and single-blind design, which provides rigorous evidence for a virtual reality (VR) intervention effect.
- The intervention has a strong ecological validity as it is done in the participants’ natural context (i.e. school) and the assessment involves students, family, and school.
- The gamified VR design is innovative and engaging, and there is an equivalent active-based VR control condition.
- One limitation is there is no follow-up assessment and no double-blind experimental design due to the nature of the study.
- A second limitation is the potential drop-out rates due the preventive nature of the interventions.

## INTRODUCTION

### Background and rationale

Previous research has shown that anxiety and depression symptoms are common in childhood and adolescence. Although the estimated prevalence in the young population varies considerably, most studies agree that anxiety disorders have currently become the most common disorders in children and adolescents (Sandín, Chorot & Valiente, 2018). The incidence of anxiety and mood disorders in adolescents is around 25% and 10 %, respectively, exceeding externalizing disorders (Avenevoli, 2012). Likewise, the negative consequences derived from anxiety symptoms can act as precursors to other mental health problems. For example, it has been observed that throughout the life cycle (including early ages such as childhood and adolescence), there is a high concurrence of anxiety disorders and mood disorders. Previous research indicates that more than 50% of people who have an anxiety disorder may develop a depressive disorder in the future and vice versa, placing the coexistence of both disorders between 40% and 80% (Wittchen et al., 2011). Even more importantly, symptoms of depression and anxiety that do not meet diagnostic criteria still contribute to considerable impairment and may develop into clinical conditions (González-Tejera et al., 2005; Comer et al., 2012).

Preventive transdiagnostic interventions for these symptoms in children and adolescents are important in reducing the development of disorders later in life, and emotion regulation (ER) is one potentially relevant factor to consider. Adjusting emotions according to environmental stimuli entails a complex and ongoing interplay of actions, cognitive processes, and physiological changes (Gross, 2015). ER involves the modification of the experience and expression of those emotions, via both automatic and controlled/delivered cognitive processes (Mauss, Bunge, & Gross, 2007; Strack & Deutsch, 2004). Deliberate processes are characterized by their need for attentional resources, their intentional nature, and their goal-driven approach. While automatic processes are triggered by mere exposure to sensory inputs, leading to the activation of pre-existing knowledge structures like schemas, scripts, or concepts, which in turn influence various psychological functions (Mauss, Bunge, & Gross, 2007). Some of these processes require attentional control, cognitive reappraisal, response modulation, emotion understanding, and working memory (Gross, 2015). Individuals with emotional dysregulation show difficulties to deliberately focus and shift attention, and to inhibit or activate behavior as appropriate to modulate their emotional states and behaviors (Compas et al., 2017). It is argued that to understand causes and mechanisms of mental disorders, many clinical symptoms can be viewed as being caused by emotionally dysregulated processes (Compas et al., 2017). The inability to effectively manage negative emotions has been linked to anxiety and mood disorders, suicidality, substance use disorders, eating disorders, post-traumatic stress disorder, and personality disorders, such as borderline personality disorder (Compas et al., 2017).

Impairments in executive functions (EF) play a critical role in the development and regulation of emotions in children and adolescents (Ochsner & Gross 2008; Lantrip et al. 2015). Processes such as working memory, inhibition, and cognitive flexibility are thought to be essential for successful ER (Lantrip et al. 2016). Research has demonstrated that children and adolescents with better EF skills are better able to regulate their emotions in response to stressful situations and exhibit fewer internalizing symptoms such as anxiety and depression (Lantrip et al. 2016). Moreover, a growing body of evidence suggests that ER and EF are closely intertwined and mutually reinforcing (Howard et al., 2021; Bartholomew, Heller & Miller, 2021). For example, inhibitory control, an essential EF skill, enables individuals to suppress maladaptive emotional responses and promote more constructive coping strategies (Camuñas et al., 2022). Even if current EF training interventions to improve ER in children and adolescents are limited, preliminary evidence suggests that boosting EF skills may prevent emotion dysregulation symptoms in this population (Rothbart & Posner, 2006).

Early detection, monitoring and preventive transdiagnostic interventions for emotion dysregulation symptoms in children and adolescents are important in reducing the development of disorders later in life (Loevaas et al., 2018). Promoting ER skills can therefore serve as a protective factor against the development of mental health disorders in children, while emotion dysregulation symptoms can increase the risk for these disorders (Chaplin et al., 2005). To do so, it is crucial to understand better the interplay between EF and ER in the context of child and adolescent mental health. Preventive early intervention strategies targeting these underlying mechanisms may mitigate the risk of developing advanced clinical disorders, such as anxiety and depression, later in life. Even if current EF training interventions to improve ER in children and adolescents are limited, preliminary evidence suggests that improving EF skills may enhance ER abilities in this population (Beloe & Derakshan, 2019; Jahn et al., 2021). Further research is needed to develop and evaluate the efficacy of such interventions, as well as to identify potential moderators and mediators of treatment outcomes, to inform more tailored and effective preventive strategies.

Immersive Virtual Reality (IVR) technology has exhibited potential to augment traditional cognitive training interventions. Several studies have concentrated on the effectiveness and applicability of IVR cognitive-based interventions. Some studies have explored IVR exergames, or games that require physical movement (Yen & Chiu, 2021; Li et al., 2016; Cugusi et al., 2020). Recent systematic review and meta-analysis reported significant gains in global cognition, memory, and a decrease in depressive symptomatology among adults utilizing IVR exergames (Li et al., 2016; Cugusi et al., 2020). Other systematic reviews have shown that IVR cognitive training is a valuable tool in treating neurocognitive disorders, with students reporting significant improvements in cognition (e.g., memory and dual-tasking) and psychological well-being (Riva et al., 2020; Moreno et al., 2019). Similar results were also observed in a systematic review involving individuals at risk of cognitive decline (Coyle et al., 2015).

The use of immersive IVR can boost the effectiveness and accessibility of cognitive training in children and adolescents. Some systematic reviews in the field have highlighted its effectiveness for improving cognitive deficits in children with attention deficit hyperactivity disorder (Romero-Ayuso et al., 2021). These immersive environments provide engaging, contextually rich stimuli, promoting neuroplasticity and potentially enhancing cognitive function (Zhu et al., 2021). And gamification parameters can increase engagement and motivation in cognitive training, while personalized and adaptive training programs can improve training effectiveness (Zhu et al., 2021; Brunner et al., 2017; Jahn et al., 2021). IVR offers a more ecological and realistic cognitive rehabilitation by creating simulations of real-world scenarios that can enhance the transfer of training effects to real-life situations (Brunner et al., 2017; Jahn et al., 2021). IVR-based programs may benefit from motivational factors, reducing attrition rates and encouraging greater motivation (Brunner et al., 2017). Furthermore, IVR creates a sense of presence with environments that simulate both internal sensory stimuli, mirroring the user’s physical sensations, and external environmental stimuli, reflecting the context of the virtual world (Cummings & Bailenson, 2015). As a result, users can explore and manipulate their environment in a self-regulated manner, enhancing their learning experiences. Through this predictive representation, IVR contributes to the improvement of movements, actions, and emotional responses, adapting them to the virtual context (Rizzo et al., 2009). This study is driven by the need to prevent the development of mental health disorders, such as anxiety and depression disorders in children and adolescents through the promotion of ER. In this research protocol, we propose to use IVR-based intervention that consists of cognitive exercises/games that have been developed based on validated neuropsychological principles and adaptive difficulty (Brugada-Ramentol et al., 2022). The intervention will target automatic cognitive processes associated with emotion dysregulation and EF, including working memory, inhibition, and cognitive flexibility.

Our **main aim** is to assess the effect of IVR-based cognitive training in children and adolescents for improving emotion dysregulation, ER strategies, and mental health status (such as anxiety and depressive symptoms), as primary measures. And cognitive functions (such as, EF, selective and sustained attention, and impulsivity) and academic performance (i.e., school grades), as secondary measures.

We anticipate that participants enrolled in the experimental IVR intervention will show significantly higher post-intervention improvements in levels of emotion dysregulation symptoms, ER skills, anxiety and depressive symptoms, EF (e.g., working memory, cognitive flexibility, inhibition control), self-efficacy, selective and sustained attention, processing speed, rumination and school grades, as compared to an active control group. We also anticipate that these improvements (i.e., significant group differences in primary and secondary measures) will be significantly greater among participants at risk for developing anxiety and depressive symptoms compared to those at low risk. As **secondary objectives** of our research study, we aim to gain valuable insights into the psychological factors (parental style or self-efficacy levels) and sociodemographic factors (such as age or gender) that may influence the effectiveness of the interventions and the magnitude of change in primary and secondary measures. Lastly, usability-based parameters (such as, the self-reported enjoyment and immersion levels, after each session), and cybersickness symptoms and general usability, at the end of the intervention.

## METHODS AND ANALYSIS

### Study design

This is a longitudinal, parallel, single-blind, randomized controlled pilot trial. The first group (experimental group) will receive the gamified IVR program (Enhance IVR by VirtuLeaps company), while the second group (active control group) will receive a comparable IVR relaxation experience with the same weekly frequency, session duration, and intervention length (see figure 1). This study will adhere to CONSORT guidelines and, it has been registered on clinicaltrials.gov (NCT06236919). The findings from this pilot-randomized controlled study will inform the design and implementation of future larger-scale randomized controlled trials.

**Figure 1.**
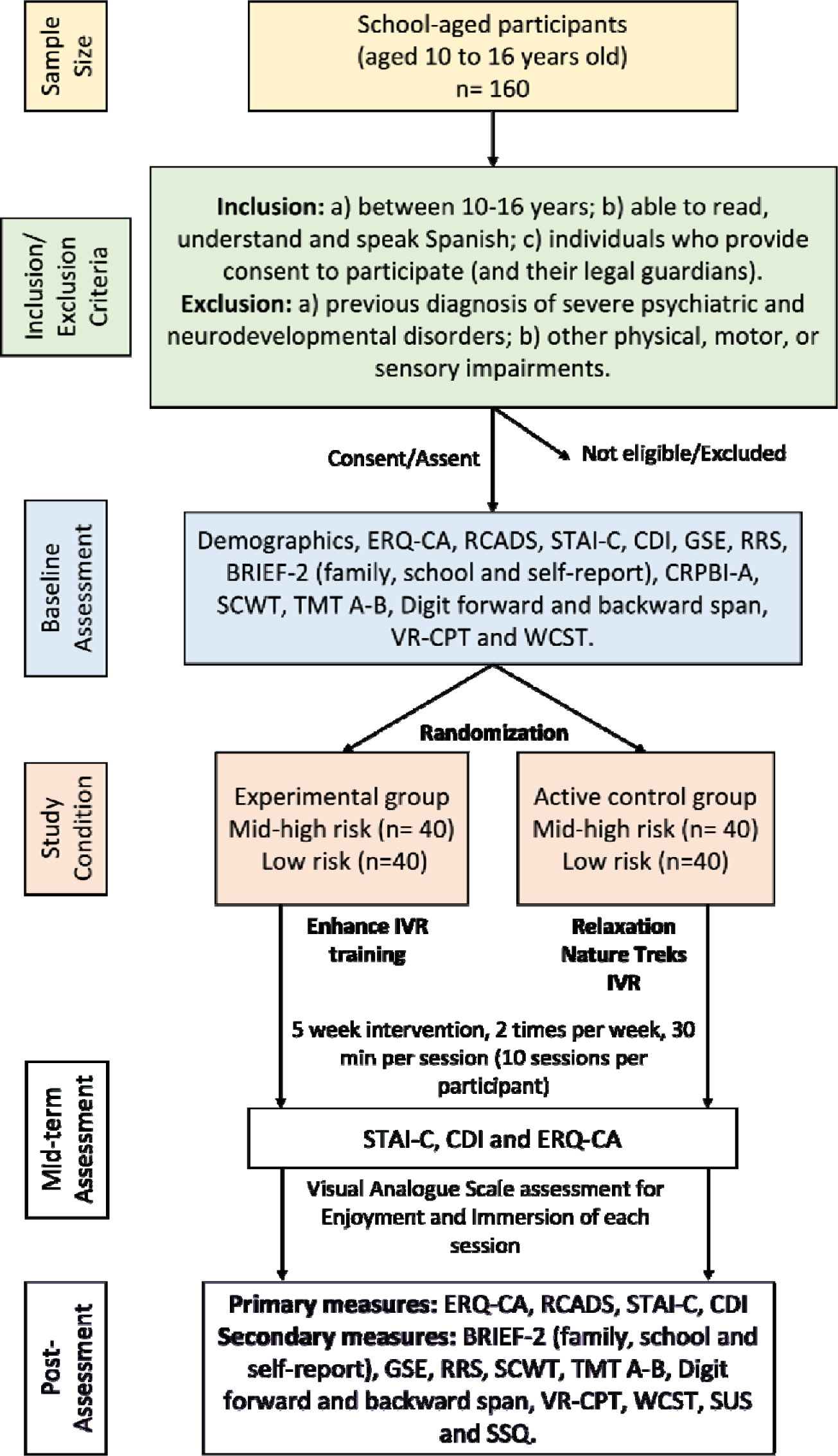
Study flow chart (CONSORT Diagram). ERQ-CA, Emotion Regulation Questionnaire for Children and Adolescents; RCADS, Revised Child Anxiety and Depression Scale; STAIC, State-Trait Anxiety Inventory for Children; CDI, Chidren’s Depression Inventory; GSE, General Self-Efficacy Scale; RRS, Ruminative Responses Scale; BRIEF-2, Behavior Rating Inventory of Executive Function 2; CRPBI-A, Child Report on Parental Behavior Inventory Abbreviated; SCWT, Stroop Color and Word Test; TMT A-B, Trail Making Test; VR-CPT, IVR-version of the Continuous Performance Test; WCST, Wisconsin Card Sorting Test; SUS, System Usabilty Scale; SSQ, Simulator Sickness Questionnaire.

### Sample

Spanish school-aged participants (aged 10 to 16 years old), whose parents and legal tutors agree to participate, will be recruited in the study. Participants will be recruited from private or public schools located in or near the metropolitan area of Barcelona, Catalonia.

#### Inclusion criteria

Participants will be included based on the following criteria: (a) between the ages of 10 years and 16 years; (b) able to read, understand and speak Spanish; and c) individuals who provide consent to participate (and their legal guardians).

#### Exclusion criteria

Participants will be excluded if they have: (a) a previous diagnosis of severe psychiatric (e.g., maniac, or psychotic symptoms) and neurodevelopmental disorders (i.e., severe autism spectrum disorders, intellectual disabilities, etc); (b) other physical, motor, or sensory impairments that could interfere with the examination or the IVR program. Participants will also be excluded if they cannot understand Spanish.

### Sample size calculation

To determine the necessary sample size for evaluating the study hypothesis through mixed between-within subject design, an a priori power analysis was conducted using *Gpower* 3.1 software. The analysis was based on similar research (Cunha et al., 2023) and aimed to achieve an 80% power for detecting a medium effect size (*partial* η*^2^* = .06), with a significance criterion of α = .05. The results indicate that a minimum sample size of *N* = 134 participants is required to achieve the desired statistical power. Furthemore, based on similar preventive studies in school-aged populations, we anticipate a 20% drop-out rate, resulting in a recruitment of 160 participants (80 participants allocated to each group).

### Procedure and randomization

Potential participants and their families will be informed about the study through a series of presentations at schools involving teachers, students, and families alike. A research team member and a school counselor will jointly screen potential participants to determine whether they meet the study’s inclusion and exclusion criteria. Once participants have agreed to participate in the study and if they meet the eligibility criteria, their legal guardians will be required to sign the informed consent form.

Next, participants will undergo a baseline assessment that includes several measures related to mental health, ER, executive functioning, and cognitive performance. Specifically, the baseline assessment battery will include sociodemographic information, the Emotion Regulation Questionnaire for Children and Adolescents (ERQ-CA, Gómez-Ortiz et al., 2016), Revised Child Anxiety and Depression Scale (RCADS-47, Sandín et al., 2009), the State-Trait Anxiety Inventory for Children (STAI-C, Spielberger, 1973), Children’s Depression Inventory (CDI, Kovacs, 1985), the General Self-Efficacy Scale (GSE, Herrero et al., 2014), Ruminative Responses Scale (RRS, Treynor et al., 2003), Behavior Rating Inventory of Executive Function 2 (BRIEF-2 self-report, family and school-based tests, Gioia et al., 2015), and the Child Report on Parental Behavior Inventory Abbreviated (CRPBI-A, Samper et al., 2006). The baseline assessment will include also a neuropsychological assessment, with the Stroop Color and Word test (SCWT, Rognoni et al., 2013), Trail Making Test (TMT A-B, Rivera & Arango-Lasprilla, 2017), Digit forward and backward span (Rivera & Arango-Lasprilla, 2017), IVR-version of the continuous performance test (IVR-CPT, Cunha et al., 2023), and the Wisconsin Card Sorting Test (WCST, Rivera & Arango-Lasprilla, 2017). All those measures will be answered by the students, except for the BRIEF that will be also answered by their school tutors and their families. Spanish-validated versions will be used for these measures for children and adolescents. Furthermore, the most recently available school grades (with scores ranging from 0 to 10) will also be registered as an index of academic performance.

In order to control for potential moderating effects of previous subclinical or clinical anxiety/depressive symptoms, all participants will first be stratified based on their anxiety and depressive risk levels. This stratification will use the RCADS-47 (Sandin et al., 2009) cut-off transformed scales for and internalizing symptoms (i.e., anxiety and depression symptoms), dividing participants into low vs mid/high-risk categories. Following stratification, participants will be randomly allocated to one of two groups— experimental or control—utilizing a basic randomization software (Robust Randomization App: RRApp; Clinical Research APPS, 2017). To ensure the integrity of the study’s simple-blind design, all outcome assessors will be systematically blinded to group allocation. This will be achieved by employing encrypted codes for group assignment, which will not be disclosed to assessors until completion of the assessments. However, due to the intrinsic characteristics of the intervention, it will be impossible to blind participants and care providers to the group allocation. To mitigate the potential biases, care providers will be trained in standardized procedures to ensure uniformity in their interactions with all participants, irrespective of group assignment. Additionally, participants will be informed about the importance of not disclosing their group status to outcome assessors, to preserve the integrity of the blinding process as much as possible.

After each VR session, participants will be assessed about the immersion and enjoyment levels they experienced while they are exposed to the IVR environment (both at the experimental and control conditions). Visual analogue scales (VAS) ranging from 0 (none) to 100 (absolutely) will be asked to each participant to obtain post-session reports. Both questions will be asked orally after completion of all IVR tasks of the session. Furthemore, there will be a mid-intervention assessment of ER, and anxiety and depressive symptoms, conducted thorugh the ERQ-CA, STAIC-S, and CDI questionnaires, respectively.

After the 5-week intervention period an assessor will re-administer the assessment battery, as in the baseline. Additionally, we will include the System Usability Scale (Sevilla-Gonzalez et al., 2020), and the Simulator Sickness Questionnaire (Campo-Prieto et al., 2021) to assess for post-intervention general usability and cybersickness symptoms. These questionnaires will be only answered by the students. All questionnaires will be administered through paper-based versions. Lastly, the most available school grades (with scores ranging from 0 to 10) will be registered again as an index of post-intervention academic performance.

### Interventions

Both the experimental group and active control group interventions will last 5 weeks, 2 times a week, 30 minutes (10 sessions per participant), with intervention parameters based on similar research in this area (Jahn et al., 2021; Cummings & Bailenson, 2015; Rizzo et al., 2009; Brugada-Ramentol et al., 2022). Before starting both intervention, each session will include a group-based psychoeducational presentation of 10 minutes of duration in which concepts related to emotions and emotional regulation will be worked on. After that, each participant will be provided with a single IVR headset, and the intervention will be delivered as a face-to-face activity in small groups of up to seven participants per session, who will be supervised by a master student, and an expert in mental health (PhD level). Study completion will be defined as participants attending 80% or more of the scheduled sessions.

### Experimental intervention

The experimental group will engage in a subset of six games developed based on the latest research in cognitive psychology and neuroscience and are aimed at improving specific cognitive and emotional functions (**Figure 2**). The first game (pizza builder) enhances attention, planification and flexibility through a dual-task paradigm in which the participant has to assemble multiple pizzas according to continuously incoming and changing orders; The second game (Maestro) enhances working memory through a n-back task in which the participant has to memorize light patterns and report them when the patterns repeat; The third game (Assembly) enhances processing speed and inhibitory control through a TMT-A task in which the participant has to select, one by one, gears from a group in ascending size; The fourth game (Memory Wall) enhances memory through a visual patterns task in which the participant has to memorize and reproduce a pattern of cubes; The fifth game (React) game enhances cognitive flexibility and inhibitory control through a WCST and Stroop task in which the participant has to throw objects into portals according to their shape or color; and the last game, Odd Egg, enhances problem solving and deductive reasoning through a category shifting task.

**Figure 2.**
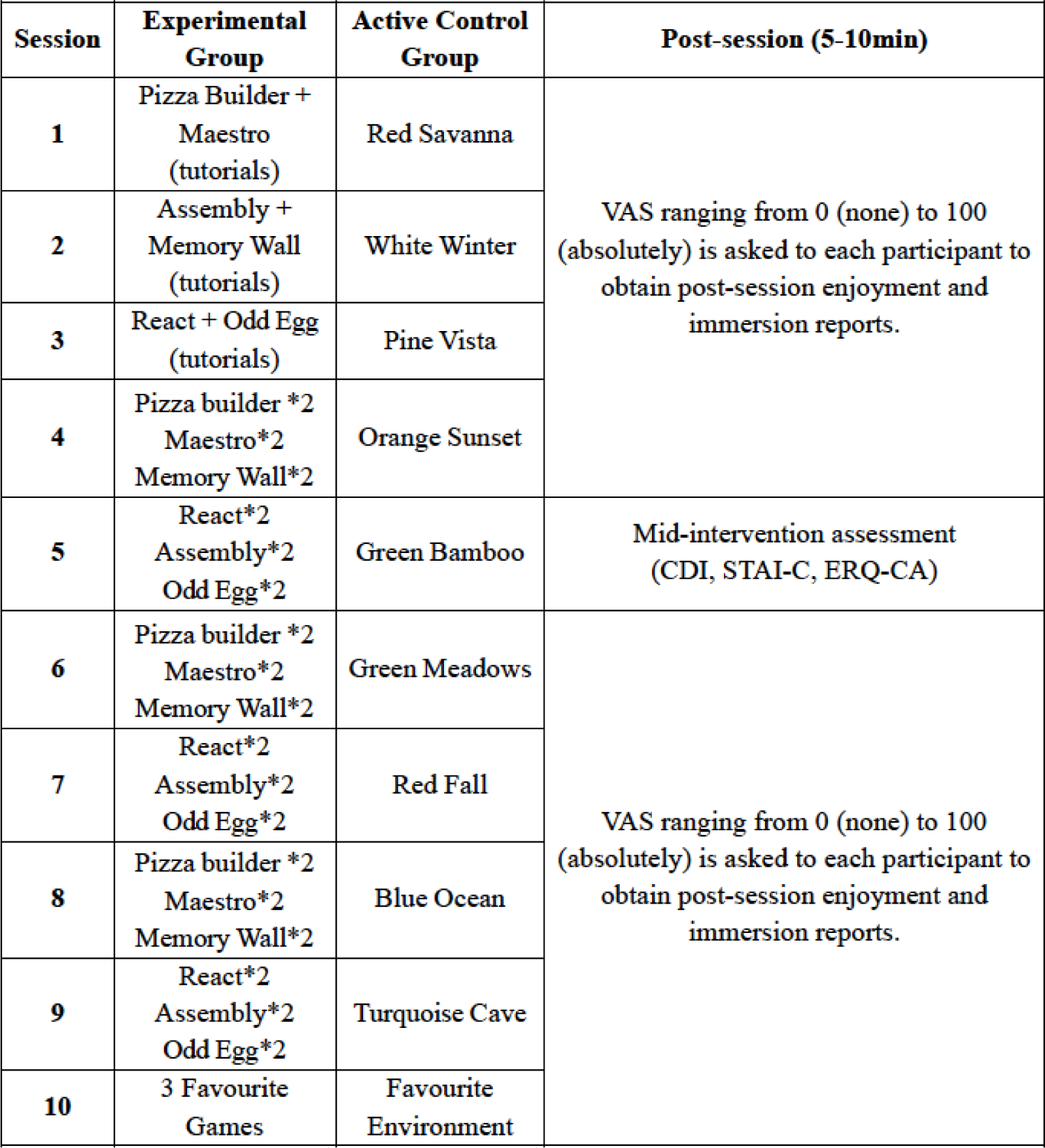
Visual scheme about e-Emotio project planification. VAS, Visual Analogue Scale.

Each gaming journey commences with a benchmark session, established to gauge the user’s initial performance level. Subsequent sessions then resume from the user’s most recent competency level, allowing for personalized progression (Brugada-Ramentol et al., 2022). The difficulty within the Enhance IVR platform is variable, contingent upon the specific game, and fine-tuned by modifying pertinent parameters for each activity. Lastly, progress tracking is facilitated through the Enhance IVR Performance Index (EPI), a calculated aggregate of performance across all cognitive domains and individual game scores.

### Active control intervention

The active control group will engage with the commercially available Nature-Trek VR software (Greener games company, USA), a sophisticated virtual reality (VR) application that simulates natural environments (**Figure 2**). These environments range from tropical islands and dense forests to serene lakes and snow-covered landscapes. Over the course of the intervention, participants will experience ten unique nature inspired IVR settings, with each session featuring a distinct environment (refer to Figure 3 for specific content of each session). The intervention protocol within each IVR session comprises three structured stages:

### VR Tailor Stage (Duration: up to 5 minutes)

Participants will be provided with an interactive opportunity to personalize their virtual environment. Through a user-friendly interface, they will be able to introduce new elements such as boulders, trees, vegetation, and swarms of butterflies, as well as alter the weather and time of day. This stage will also allow participants to navigate through the environment using their preferred method of locomotion (either fixed-point teleportation or arm-swing free locomotion) to select their desired location for the subsequent stages of the session.

### Deep Breathing Training Stage (Duration: up to 4 minutes)

Based on previous evidence from prior research indicating the effectiveness of breathing exercises in ER (Stromberg et al., 2015; Broderick, 2009) and reducing anxiety, depression, and stress (Jerath et al., 2015; Ma et al., 2017; Magnon, Dutheil & Vallet, 2021), this stage introduces a structured relaxation technique (i.e., deep breathing technique). Participants will engage with a visual representation of a lotus that synchronizes with the optimal rhythm for diaphragmatic breathing—encouraging deep breaths, belly expansion, and a slower respiration rate to enhance gas exchange and relaxation.

### Immerse Stage (Duration: 6 minutes)

In the final stage, participants are encouraged to relax fully and immerse themselves in the virtual setting without specific directives. They are advised to explore and identify personal relaxation strategies that work best for them. No further instructions will be given in this last stage that will have a fixed duration of 6 minutes as it has been done in previous studies with VR settings (Browning et al., 2020).

### Hardware

The stand-alone, Meta Quest 3 head-mounted display (HMD), introduced in October 2023, will be used in both groups. This device, succeeding the Meta Quest 2, signifies a notable evolution in VR technology, and incorporates the Snapdragon XR2 Gen 2 chipset. The Meta Quest 3 is characterized by its high-resolution display, offering 2064×2208 pixels per eye, coupled with a refresh rate of 120 Hz and a horizontal field of view extending to 110 degrees. These features collectively facilitate an immersive user experience devoid of external base stations for tracking. Complementing the technical specifications, the device is furnished with two 6DoF (Degrees of Freedom) controllers, enabling precise and intuitive user interaction within the virtual environment.

### Statistical analysis

Analyzes will be conducted with the SPSS and R studio software. Baseline sociodemographic characteristics of all participants will be described for continuous and categorical data. To assess the primary study aim, three-way mixed ANOVAs, with group condition (experimental vs control) and anxiety/depressive risk levels (mid/high vs low) as a between subject factors and assessment time (pre vs post assessment) as within-subject factor, will be conducted for all the primary and secondary measures assessed on the study. Post hoc corrections will be applied for multiple comparisons (Bonferroni corrections). The analyses will be Intention to Treat and per protocol. The critical level of statistical significance will be set at α = .05. Effect size will be calculated using partial eta squared (*partial* η*^2^*). Graphs and descriptive statistics will be used to examine assumptions of the tests.

Secondly, the magnitude of change of the interventions (or intervention gains) will be computed by subtracting pre-post differences for both primary and secondary measures of the study. Then, Pearson correlation coefficients will be used to examine the relationships between potential intervention moderators (such as, age, gender, baseline self-efficacy levels, and baseline educational parental style) and the pre-post differences in primary and secondary measures of the study. This approach allows us to explore how these moderating factors may influence the effect of both interventions. Pearson correlations assume that the data is normally distributed and that the relationships between variables are linear. If these assumptions are not met, we will consider using non-parametric alternatives, such as Spearman’s rank-order correlations.

To assess the impact on immersion and enjoyment levels experienced over the course of the interventions, we will employ a mixed-model analysis to examine differences in VASs collected following each IVR session. This analytical approach will allow us to understand the variations in participant responses over time (i.e., in 10 sessions) and between both groups. Additionally, we aim to evaluate differences in usability and the occurrence of cybersickness symptoms because of the intervention. Thus, we will conduct independent-sample t-tests (or the non-parametric alternative in case of not meeting the tests assumptions), for assessing group differences on usability parameters and cybersickness symptoms after the completion of the interventions.

### Patient and Public Involvement

Participants were not involved in the design, or conduct, or reporting, or dissemination plans of our research.

## ETHICS, SAFETY AND DISSEMINATION

### Ethics

Ethical approval of the study protocol and the informed consent forms was received and approved by the Ethics Committee of the International University of Catalonia (PSI-2023-04). Participants will be fully informed with written consent and can always choose to stop participating in this study. This protocol includes clear delineation of the protocol version, and no major protocol modifications are anticipated during the duration of this study.

### Safety

All data collected are for research purposes only, and data will be kept in strict confidence. No information will be given to anyone without permission from the participants and legal guardians. Confidentiality will be ensured by use of identification codes. All data, whether generated in the study, will be identified with a randomly generated identification code unique to the subject. The information collected for this research study will be accessible to authorized persons. Authorized persons include the principal investigator, research team members, representatives of Universitat Internacional de Catalunya; and representatives of other agencies when required by law. Paper research records and hard copy data will be kept in a secure location (in locked file cabinets, within a locked office), and electronic research files and data will be kept on password protected and encrypted computers. Furthermore, research files and its contents will only be labeled with a code, and a master list of codes and identities will be maintained in a secure location apart from the coded data. Participant identifiers will be stored separately from the coded participant data. Data gathered in this study (including sociodemographic data and all psychological and neuropsychological measures) will not be transferred to the virtual reality providers. At the conclusion of this study the researchers may publish their findings. Information will be presented in summary format, and participants will not be identified in any publications or presentations.

The research intervention will be conducted in private offices or classrooms in schools. The collection of information about participants will be limited to the amount necessary to achieve the aims of the research, so that no unneeded information will be collected.

Periodically, data and documentation will be monitored as each participant completes the study. The following data and documentation will be monitored by the two investigators, through the study: a) Periodic review and confirmation of participant eligibility. b) Periodic review of informed consent documentation. c) Periodic review of the transfer/transcription of data from the original source to the research record.

The stopping rules for this study involve: (1) the intervention is associated with adverse effect that call into question the safety of the intervention; (2) difficulty in study recruitment or retention will significantly impact the ability to evaluate the study endpoints; (3) any new information becomes available during the trial that necessitates stopping the trial; or (4) other situations occur that might warrant stopping the trial.

Adverse events (AE) reports will not include subject- or group-identifiable material. Each report will only include the identification code. AEs will be labeled according to severity, which is based on their impact on the student. An AE will be termed “mild” if it does not have a major impact on the student, “moderate” if it causes the student some minor inconvenience, and “severe” if it causes a substantial disruption to the student’s well-being. AEs will be categorized according to the likelihood that they are related to the study intervention. Specifically, they will be labeled unrelated, definitely related, probably related, or possibly related to the study intervention.

### Dissemination

Findings and results will be disseminated nationally and internationally through peer-reviewed journals, reports, scientific conferences and at general public events. Participants’ parents and teachers may receive trial results if interested. All co-authors are eligible to participate in dissemination events and it is not planned to use professional writers in this results dissemination.

### Future potential and implications of the current study

The preliminary findings from this research have the potential to contribute significantly to the development of structured and larger-size effective prevention strategies to reduce mental health disorders in children and adolescents. We anticipate an improvement on those aspects of mental health in younger populations can have a profound impact on their overall well-being, as reducing anxiety and depressive symptoms and promoting better ER can lead to improved academic performance, social functioning, and resilience to future stressors (Wexler et al., 2016). From an educational perspective, the use of IVR cognitive training can help students develop problem-solving skills and the ability to manage their emotions in challenging academic situations as well as enhance their academic performance by improving EF, attention, concentration, and memory (Hajal & Paley, 2020). Socially, the intervention can improve social skills, empathy, and social interaction, leading to positive social outcomes for the individual and their peers (Carreon et al., 2020).

The use of immersive technological solutions, as IVR, represents a particular opportunity for improving school-based preventive interventions for several reasons. Firstly, IVR provides a safe and controlled environment in which children and adolescents can practice skills in a realistic and immersive way (Dix et al., 2011). By simulating real-life scenarios, IVR cognitive training can help individuals develop and refine their ER skills, such as identifying and managing negative emotions and coping with stressors (Schwean & Rodger, 2013). Also, cognitive training through IVR technology can be tailored to meet the specific needs of everyone, as it allows for individualized feedback and adaptive difficulty levels (Brugada-Ramentol et al., 2022). This is particularly important for school-aged children and adolescents, who may have different levels of emotional regulation skills or respond differently to different types of stimuli. By adapting the IVR cognitive training to everyone’s cognitive profile, we expect that this intervention will be more effective in improving ER skills and reducing symptoms of anxiety and depression. Finally, the use of IVR technology in this preventive intervention can be engaging and enjoyable for children and adolescents, which can increase motivation and participation, increasing the likelihood that they will complete the program and benefit from the intervention. This is particularly important for a preventive intervention, as maintaining engagement and participation over time can be a challenge in school-based settings (Fazel et al., 2014).

Considering these opportunities, this project will test an evidence-based, accessible, and feasible IVR solution adapted to children and adolescents that can be cost-effective (i.e., by using stand-alone IVR headsets) and easily disseminated in schools. The research team will provide unique clinical, research, educational, technological, and translational expertise to execute this proposal.

## Data Availability

The data sets generated and/or analyzed during this study are available from the corresponding author upon reasonable request.

## Acknowledgements

This study was funded and supported by a NARSAD Young Investigator Grant from the Brain & Behavior Research Foundation (ID: 31607). We also express our sincere gratitude to Mr. Amir Bozorgzadeh, and to Dr. Bebiana Moura, Head of Partnerships from Virtuleaps, for his pivotal technical support to help us design the research protocol of Enhance IVR virtual games. Their expertise and dedication, along with the invaluable assistance of the VirtuLeaps team, will be instrumental in tackling intricate technical issues before and during the recruitment.

## Authors’ contributions

BPG and MGF wrote the first draft of the research protocol, ACM, AGC and KA refined and perfected the first draft into the current manuscript. ACM, AGC, KA and JRR will be conducting the project launch, recruitment of participants, baseline data collection, study sessions and post-assessment data collection and site-management of the study. MFC, MGS, and MGF participated on the initial research design and manuscript revision. BPG is the principal investigator of this project in the management and leadership of the research project, with tasks including research design, intervention design and implementation, eligibility assessments, manuscript revision, project management and supervision. All authors have read and agreed to the published version of the manuscript.

## Copyright

I, the Submitting Author has the right to grant and does grant on behalf of all authors of the Work (as defined in the <u>author licence</u>), an exclusive licence and/or a non-exclusive licence for contributions from authors who are: i) UK Crown employees; ii) where BMJ has agreed a CC-BY licence shall apply, and/or iii) in accordance with the terms applicable for US Federal Government officers or employees acting as part of their official duties; on a worldwide, perpetual, irrevocable, royalty-free basis to BMJ Publishing Group Ltd (“BMJ”) its licensees. The Submitting Author accepts and understands that any supply made under these terms is made by BMJ to the Submitting Author unless you are acting as an employee on behalf of your employer or a postgraduate student of an affiliated institution which is paying any applicable article publishing charge (“APC”) for Open Access articles. Where the Submitting Author wishes to make the Work available on an Open Access basis (and intends to pay the relevant APC), the terms of reuse of such Open Access shall be governed by a Creative Commons licence – details of these licences and which licence will apply to this Work are set out in our licence referred to above.

